# Clinical outcomes in vaccinated individuals hospitalized with Delta variant of SARS-CoV-2

**DOI:** 10.1101/2021.07.13.21260417

**Authors:** V Jagadeesh Kumar, Divya Tej Sowpati, Apoorva Munigela, Sofia Banu, Archana Bharadwaj Siva, M Sasikala, Chandrasekhar Nutalapati, Anand Kulkarni, Payel Mukherjee, Lamuk Zaveri, CCMB COVID-19 Team, AIG Hospitals COVID-19 Vaccine study Team, Karthik Bharadwaj Tallapaka, D Nageshwar Reddy

**Author notes:** Corresponding authors Dr. D Nageshwar Reddy, MD, DM, FAMS, FRCP, FASGE, FACG, MWGO, FAGA, FJGES, AAAS, AIG Hospitals, Mindspace Rd, Gachibowli, Hyderabad - 500032, Telangana, IN, Email -, Dr. Karthik Bharadwaj Tallapaka, MD, DNB, CSIR - Centre for Cellular and Molecular Biology, Uppal Road, Habsiguda, Hyderabad - 500007, Telangana, IN. Equal Contribution (V Jagadeesh Kumar, Divya Tej Sowpati).

## Abstract

Emerging variants of SARS-CoV-2 with increased transmissibility or immune escape have been causing large outbreaks of COVID-19 infections across the world. As most of the vaccines currently in use have been derived from viral strains circulating in the early part of the pandemic, it becomes imperative to constantly assess the efficacy of these vaccines against emerging variants. In this hospital-based cohort study, we analysed clinical profiles and outcomes of 1161 COVID-19 hospitalized patients (vaccinated with COVISHIELD (ChAdOx1) or COVAXIN (BBV-152), n = 495 and unvaccinated n = 666) in Hyderabad, India between April 24^th^ and May 31^st^ 2021. Viral genome sequencing revealed that >90% of patients in both groups were harbouring the Delta variant (Pango lineage B.1.617.2) of SARS-CoV-2. Vaccinated individuals showed higher neutralizing antibodies (545±1256 AU/ml Vs 51.1±296 AU/ml; p<0.001) and significantly decreased Ferritin (392.26 ± 448.4 ng/mL Vs 544.82 ± 641.41 ng/mL; p<0.001) and LDH (559.45 ± 324.05 U/L Vs 644.99 ± 294.03 U/L; p<0.001), when compared to the unvaccinated group. Severity of the disease (3.2% Vs 7.2%; p=0.0039) and requirement of ventilatory support (2.8% Vs 5.9%; p=0.0154) were significantly low in the vaccinated group despite the fact that these individuals had significantly higher age and risk factors. The rate of mortality was about 50% lower (2/132=1.51%) in the completely vaccinated breakthrough infections although mortality in individuals who had received a single dose was similar to the unvaccinated group (9/269=3.35% vs 23/666= 3.45%). Our results demonstrate that both COVISHIELD and COVAXIN are effective in preventing disease severity and mortality against the Delta variant in completely vaccinated hospitalized patients.

## Introduction

New variants of SARS-CoV-2 continue to emerge as the virus spreads among hosts and mutates. Some of the variants, termed variants of concern, show increased transmissibility or capacity to bypass existing immunity, and are associated with large outbreaks of COVID-19 infections across the world (1,2). The Delta variant or the lineage B.1.617.2 of SARS-CoV-2, first sampled in late 2020 in India, is a highly transmissible variant and has been associated with the large second wave of COVID-19 in India (3). The variant has now spread to more than 92 countries (4) and has become the largest circulating viral strain in the world. From the experience so far, vaccination has been the most effective public health intervention in curtailing the spread of the pandemic. As most of the vaccines currently in use have been derived from viral strains circulating in the early part of the pandemic, it becomes imperative to constantly assess their efficacy against emerging variants and make changes to the vaccine composition, as and when necessary (5). *In vitro* studies so far have revealed decreased neutralizing efficacy of the Pfizer-BioNTech vaccine (BNT162b2) and the Oxford-AstraZeneca vaccine (ChAdOx1) against the Delta variant (6). In the current study, we assessed the clinical outcomes of hospitalized COVID-19 patients vaccinated with either COVISHIELD (ChAdOx1, Serum Institute of India) or COVAXIN (BBV-152, Bharat Biotech) and compared them with unvaccinated individuals admitted to a Covid care facility between April 24^th^ 2021 and May 31^st^ 2021.

## Results and Discussion

A total of 1161 SARS-CoV-2 infected patients admitted into AIG Hospitals (a tertiary care referral hospital in Hyderabad, India) during the above mentioned time period, were enrolled in the study. Of these, 495 were vaccinated, (mean age - 58.50 ± 13.10 years) and 666 were unvaccinated (mean age - 47.20 ± 14.70 years). Patients in the vaccinated group were of significantly higher age (*p*<0.001) [Table 1], as individuals above 45 years were prioritized in the initial vaccination drive in accordance with the guidelines of the Government of India. Out of 495 vaccinated, 251 received COVISHIELD (168 single dose and 83 two doses), 203 received COVAXIN (99 single dose and 104 two doses) and in the remaining 41, the vaccine type or number of doses was not known. Significantly higher numbers of patients in the vaccinated group had comorbidities such as diabetes, hypertension and cardiovascular disease (*p*<0.001) as compared to the unvaccinated group. Viral genomes could be sequenced from the nasopharyngeal swabs obtained from 201 individuals (vaccinated: n=97, unvaccinated: n=104) during the study period. More than 90% of the individuals were infected with the Delta variant (Pango lineage B.1.617.2) in both the groups (93 in vaccinated & 94 in unvaccinated groups respectively) [Figure 1A]. This was comparable to the prevalence of the variant in the community during this period [Figure S1]. Hence, it may be safe to assume that the proportions will hold true in the remaining samples where viral genome sequencing could not be done.

**Table 1:** Clinical profiles, inflammatory markers and outcomes of vaccinated and unvaccinated SARS-CoV-2 positive patients

|  | **Unvaccinated**<br>**(n = 666)** | **Vaccinated**<br>**(n = 495)** | ***p-value*** |
| --- | --- | --- | --- |
| Age | 47.2 ± 14.7 | 58.5 ± 13.1 | < 2.2e-16 |
| Females | 239 (35.9%) | 180 (36.4%) | 0.8536 |
| Diabetes/Hypertension | 243 (36.5%) | 256 (51.7%) | 2.448e-07 |
| Cardiovascular disease | 18 (2.7%) | 37 (7.5%) | 2.21E-04 |
| Respiratory diseases | 14 (2.1%) | 8 (1.6%) | 0.6653 |
| Malignancy | 4 (0.6%) | 5 (1.0%) | 0.5078 |
| Chronic kidney disease | 15 (2.3%) | 12 (2.4%) | 0.8466 |
| Chronic liver disease | 9 (1.4%) | 5 (1.0%) | 0.7872 |
| C-Reactive Protein (mg/l) | 45.35 ± 54.43 (n = 617) | 45.74 ± 56.51 (n = 461) | 0.9083 |
| Ferritin (ng/ml) | 544.82 ± 641.41 (n = 609) | 392.26 ± 448.4 (n = 461) | 5.31E-06 |
| D-dimer (ng/ml) | 436.18 ± 739.56 (n = 617) | 438.49 ± 844.83 (n = 461) | 0.9627 |
| LDH (U/L) | 644.99 ± 294.03 (n = 538) | 559.45 ± 324.05 (n = 402) | 3.45E-05 |
| Total Leukocyte count (Cells/cu.mm) | 7087.01 ± 3590.57 (n = 615) | 7135.38 ± 3539.59 (n = 455) | 0.8262 |
| Neutrophils (Cells/cu.mm) | 5296.56 ± 3371.13 (n = 614) | 5154.92 ± 3224.98 (n = 455) | 0.4863 |
| Lymphocytes (Cells/cu.mm) | 1341.11 ± 711.73 (n = 607) | 1400.5 ± 648.52 (n = 453) | 0.1575 |
| Platelets (lakhs/cu.mm) | 2.3 ± 0.83 (n = 607) | 2.21 ± 0.75 (n = 454) | 0.0685 |
| Baseline neutralizing antibodies (AU/ml) | 51.1 ± 296 | 545 ± 1256 | 1.663e-14 |
| Severe disease/ ICU requirement at admission | 48 (7.2%) | 16 (3.2%) | 0.0039 |
| ICU requirement during the hospitalization | 31 (4.7%) | 19 (3.8%) | 0.5601 |
| Ventilatory support | 39 (5.9%) | 14 (2.8%) | 0.0153 |
| Thrombotic complications | 19 (2.9%) | 11 (2.2%) | 0.5779 |
| Acute kidney injury | 30 (4.5%) | 24 (4.8%) | 0.7801 |
| Renal Replacement Therapy | 5 (0.8%) | 6 (1.2%) | 0.5433 |
| Death | 23 (3.45%) | Single Dose: 9 (3.35%)<br>Fully Vaccinated: 2 (1.51%) | 1.0000<br>0.4086 |

**Figure 1.**
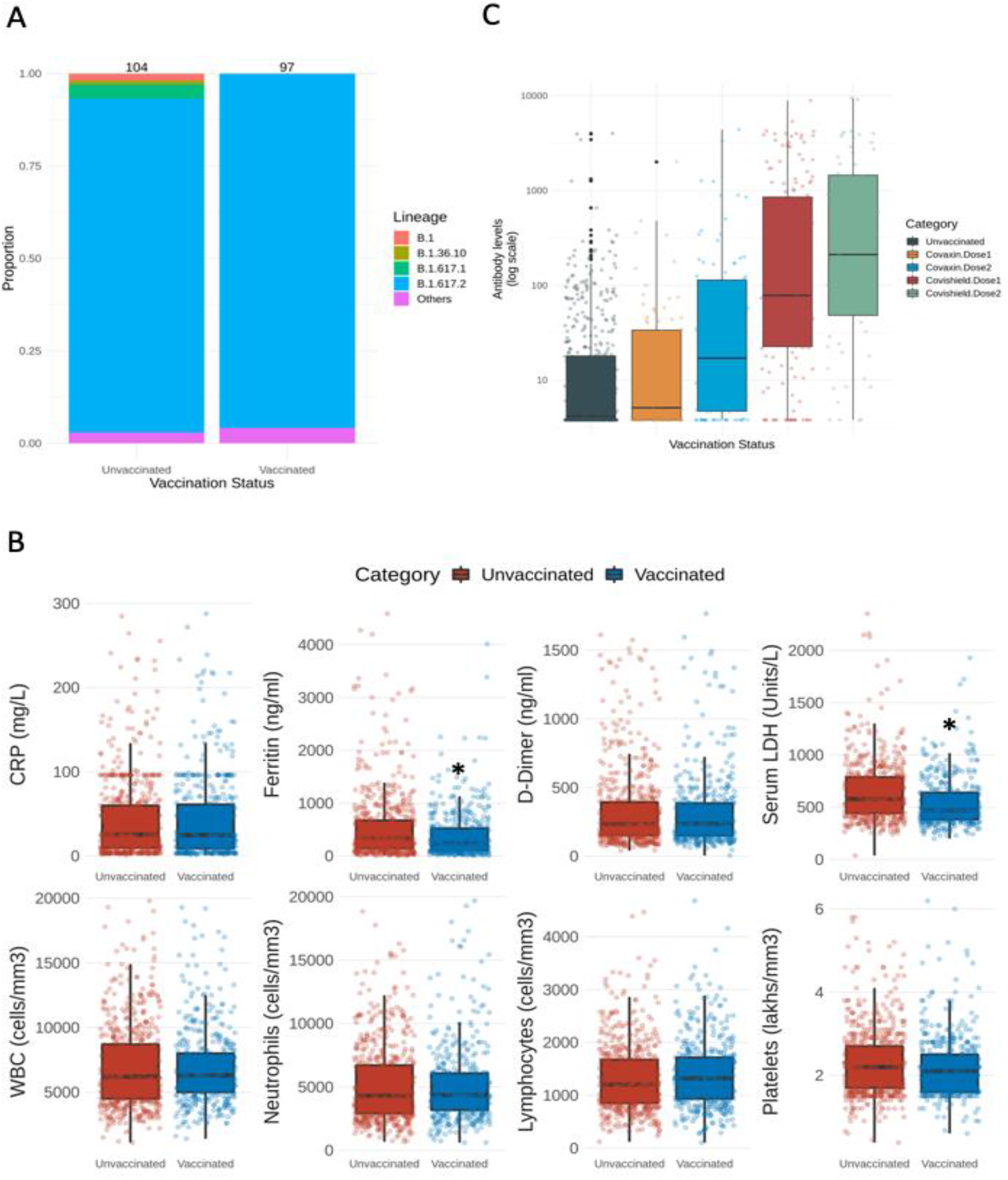
**(A)** A stacked bar plot representing the proportion of various SARS-CoV-2 lineages in vaccinated and unvaccinated individuals. Total number of genomes ina group are indicated on the top of the bar. **(B)** Inflammatory markers and Immune profiles in vaccinated and unvaccinated individuals. Significant differences are indicated with an asterisk. (**C)** Neutralizing antibody levels in vaccinated and unvaccinated individuals.

Vaccinated individuals showed significantly higher neutralizing antibodies (545±1256 AU/ml Vs 51.1±296 AU/ml; p<0.001) and lower inflammatory markers like serum Ferritin (392.26 ± 448.4 ng/mL Vs 544.82 ± 641.41 ng/mL; p<0.001) and Lactate dehydrogenase (LDH; 559.45 ± 324.05 U/L Vs 644.99 ± 294.03 U/L; p<0.001), when compared to the unvaccinated group suggesting possible early neutralization of the virus and thereby prevention of aberrant inflammatory response [Figure 1B]. A trend of increasing antibody levels and decreasing inflammatory markers was noted in individuals who received both the doses when compared with those who received a single dose [Figure 1C, Figure S2]. Individuals having higher antibody titres in the unvaccinated group were found to have IgM antibodies suggesting that these were acute infections and not reinfections or breakthrough infections.

Severity of the disease (Severity/ICU requirement at admission) was significantly low in the vaccinated group when compared to the unvaccinated (3.18% Vs 7.06%; p=0.0039). Requirement of ventilatory support was low as well (2.8% Vs 5.7%; p=0.0156). No significant differences were seen in the incidence of acute kidney injury (AKI), requirement for renal replacement therapy (RRT) or thrombotic complications between both the groups [Table 1]. Mortality was 50% lower (<u>2/132=1.51%</u>) in fully vaccinated individuals (completed 14 days after receiving second dose) hospitalised with SARS-CoV-2 infection when compared to unvaccinated individuals (<u>23/666=3.45%</u>) while it was similar (<u>9/269=3.35%</u>) in single dose vaccinated individuals. These results become even more significant in the light of higher comorbidities and age in the vaccinated group. Majority of deceased in completely and partially vaccinated individuals had no/minimal antibody response which was comparable to unvaccinated individuals. Strategies targeting these non-responders to vaccination like additional booster doses or change of vaccine type need to be explored further.

Within the vaccinated group, neutralizing antibody response and total leukocyte counts were significantly higher, and serum Ferritin levels were significantly lower in those who received COVISHIELD (both doses) than those who received COVAXIN (both doses) [Table 2]. However, no significant difference was seen in severity/mortality between both the groups. The clinical profiles of both the vaccinated groups were comparable (Table 2).

**Table 2:**
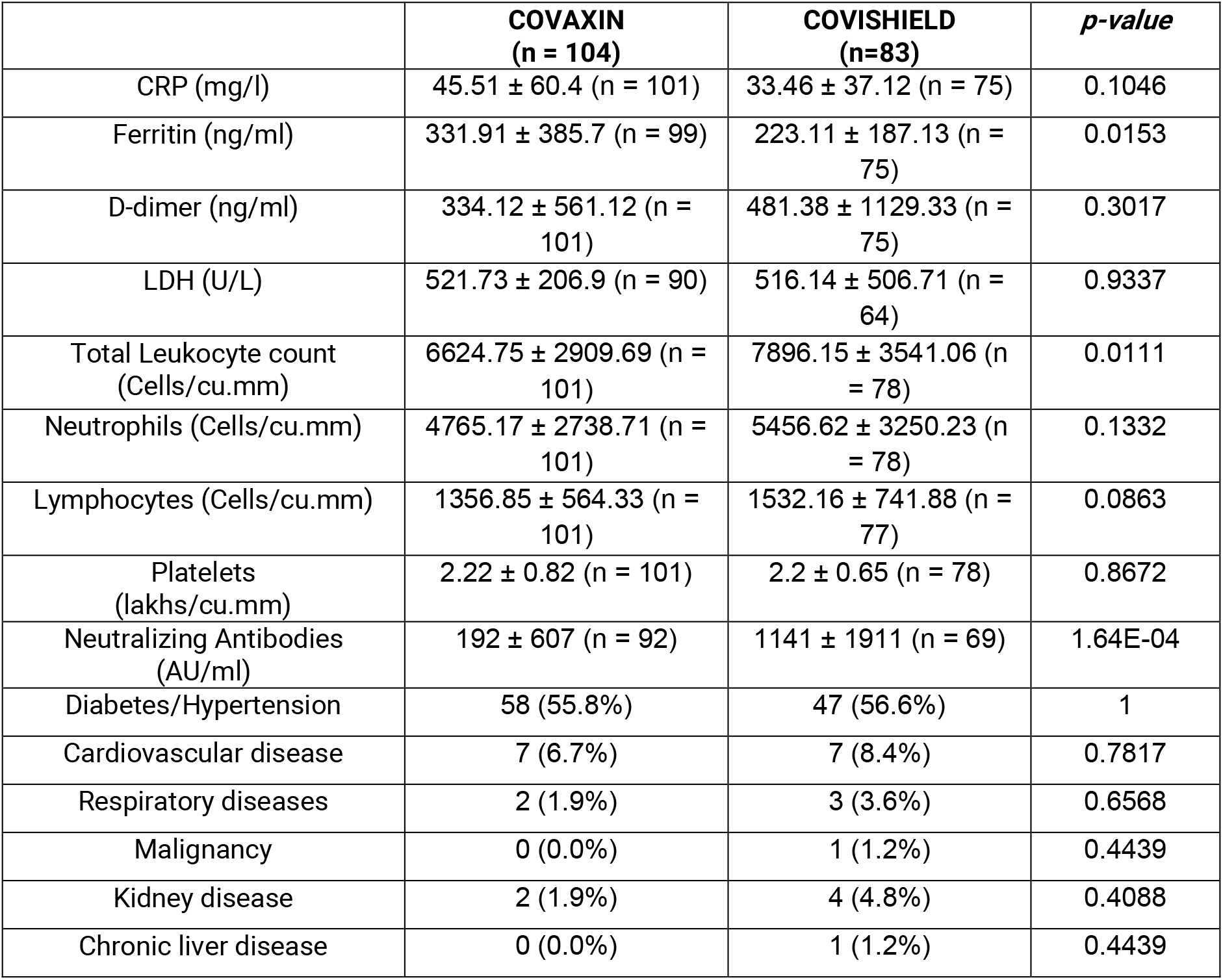
Comparison of inflammatory markers, comorbidities and outcomes between COVAXIN and COVISHIELD (both doses received)

|  | **COVAXIN**<br>**(n = 104)** | **COVISHIELD**<br>**(n=83)** | ***p-value*** |
| --- | --- | --- | --- |
| CRP (mg/l) | 45.51 ± 60.4 (n = 101) | 33.46 ± 37.12 (n = 75) | 0.1046 |
| Ferritin (ng/ml) | 331.91 ± 385.7 (n = 99) | 223.11 ± 187.13 (n = 75) | 0.0153 |
| D-dimer (ng/ml) | 334.12 ± 561.12 (n = 101) | 481.38 ± 1129.33 (n = 75) | 0.3017 |
| LDH (U/L) | 521.73 ± 206.9 (n = 90) | 516.14 ± 506.71 (n = 64) | 0.9337 |
| Total Leukocyte count (Cells/cu.mm) | 6624.75 ± 2909.69 (n = 101) | 7896.15 ± 3541.06 (n = 78) | 0.0111 |
| Neutrophils (Cells/cu.mm) | 4765.17 ± 2738.71 (n = 101) | 5456.62 ± 3250.23 (n = 78) | 0.1332 |
| Lymphocytes (Cells/cu.mm) | 1356.85 ± 564.33 (n = 101) | 1532.16 ± 741.88 (n = 77) | 0.0863 |
| Platelets (lakhs/cu.mm) | 2.22 ± 0.82 (n = 101) | 2.2 ± 0.65 (n = 78) | 0.8672 |
| Neutralizing Antibodies (AU/ml) | 192 ± 607 (n = 92) | 1141 ± 1911 (n = 69) | 1.64E-04 |
| Diabetes/Hypertension | 58 (55.8%) | 47 (56.6%) | 1 |
| Cardiovascular disease | 7 (6.7%) | 7 (8.4%) | 0.7817 |
| Respiratory diseases | 2 (1.9%) | 3 (3.6%) | 0.6568 |
| Malignancy | 0 (0.0%) | 1 (1.2%) | 0.4439 |
| Kidney disease | 2 (1.9%) | 4 (4.8%) | 0.4088 |
| Chronic liver disease | 0 (0.0%) | 1 (1.2%) | 0.4439 |

To conclude, Delta variant has been the predominant cause of breakthrough infections during the second wave in India. Our preliminary results indicate that both COVISHIELD and COVAXIN offer protection and have comparable efficacies against the Delta variant resulting in reduction of the disease severity and mortality among the hospitalized patients with two doses of vaccination. Rapid and complete vaccination of the population, therefore, remains our only hope in mitigating the deadly pandemic.

## Methods

### Patient recruitment

In this retrospective cohort study, all SARS-CoV-2 reverse transcription polymerase chain reaction (RT-PCR) positive patients admitted to AIG hospitals, Hyderabad, India between April 24^th^ 2021 and May 31^st^ 2021 were included (n=1161). The primary aim was to assess the effectiveness of COVISHIELD and COVAXIN on the clinical outcomes in hospitalized patients harbouring variant of concern B.1.617.2. Clinical data and lab results were collected and analysed as described below.

### Definitions

Patients who had taken vaccination (single/double dose) prior to contracting the infection were categorized under the vaccinated group. Those who had not taken the vaccine were categorized under unvaccinated group. Those who completed 2 weeks after the second dose were considered fully vaccinated. The disease severity was defined as per the current Indian Council Medical Research (ICMR) guidelines. According to ICMR guidelines, patients with upper respiratory tract symptoms and without dyspnea (or hypoxia) but with or without fever were categorized to have mild disease, patients with room air oxygen saturation between 90-93% to have moderate disease, and patients with room air oxygen saturation <90% to have severe disease [<u>Clinical Guidance for Management of Adult COVID-19 Patients</u> (icmr.gov.in)]. Severe disease at admission was defined as patients with SpO2 <90% or requirement of ICU care. Mortality occurring anytime during the hospital stay was considered as in-hospital mortality. Patients on anti-diabetic medications irrespective of the duration of the therapy were considered diabetic. Patients on anti-hypertensive therapy were considered hypertensive. Patients with bronchial asthma, chronic obstructive pulmonary disease, interstitial lung disease were grouped as pre-existing respiratory diseases. Patients with a history of coronary interventions or interventions for cardiac rhythm disturbances, and pre-existing cerebrovascular events were grouped under cardiovascular disease. Patients with clinically, biochemically, or histologically proven liver disease were considered to have chronic liver disease. Patients on maintenance hemodialysis or eGFR <60 for more than three months were considered to have chronic kidney disease. Patients developing thrombotic complications such as deep vein thrombosis, pulmonary thromboembolism, acute coronary syndromes, cerebrovascular accidents, and transient ischemic attack and embolism of abdominal vessels were grouped under thromboembolic complications. COVID-19 infected patients requiring mechanical ventilation or non-invasive ventilation for respiratory failure were categorized under the group requiring ventilatory support. All patients received treatment as per standard protocols and were followed for their progress during their hospital stay until discharge or death. The study was approved by the Ethics committee of AIG Hospitals & CSIR-CCMB and the informed consent was waived. All the authors had full access to the deidentified data.

### Whole genome sequencing of SARS-CoV-2

Viral RNA from COVID-19 infected patients was sequenced using COVIDSeq (Illumina, USA) according to the manufacturer’s protocol. Briefly, first strand cDNA was synthesized using random hexamers and reverse transcriptase. Viral first strand cDNA was amplified using two primer pools. Post amplification, the two pools were combined, tagmented, and purified. An amplification step was performed to add a 10 base index and sequencing adapter, and the samples were purified using paramagnetic beads to ensure selection of optimal sized fragments required for sequencing. Purified samples were quantified, normalized and sequenced on a NovaSeq 6000 (Illumina, USA) to obtain 100bp paired-end reads.

### Data processing

Base calling was performed on raw image data using bcl2fastq v2.20.0.422 (Illumina). FASTQC v0.11.9 (7) was used to assess the quality of reads. Trimmomatic was used to trim poor quality bases and adapter sequences (8). Mapping of reads to the indexed reference genome NC_045512.2 was done using HISAT2 v2.1.0 (9). Consensus sequences were generated using bcftools from the BAM file post alignment. Coverage across the genome was calculated using samtools depth. PANGO v3.0.5 (3) was used to assign lineages to the consensus sequences. Mutations in the sequences were identified using Nextclade v1.1.0 (10).

### Statistical analysis

Data was analyzed using the R statistical environment. Categorical data including gender, comorbidities, outcomes of COVID-19 is expressed as numbers (%). Continuous variables including age, biochemical variables (hemogram, LFT and serum creatinine), inflammatory variables (Ferritin, LDH, CRP, and D-dimer) are expressed as means (standard deviation [SD]). Categorical data was compared using Fisher’s exact test while continuous data was compared using Student’s t-test. All statistical tests with *p*<0.05 were considered significant. Data was visualized using the ggplot2 package in R.

## Data Availability

The data is available with the corresponding Author from AIG Hospitals and is available upon request

## Acknowledgements

This study is supported by the CSIR grant MLP0128 and intramural funds from Asian Healthcare Foundation.

## CCMB COVID19-Team

Sreenivas Ara

Sumedha Avadhanula

Himasri Bollu

Sai Krishna Jandhyala

Onkar Kulkarni

Vidhyadhari Methuku

Sai Priya Nurkurthy

Valli Undamatla

Shreekant Verma

Amareshwar Vodnapalli

## AIG Hospitals COVID-19 Vaccine study Team

Ravikanth Vishnubotla

Krishna Vemula

Jayashree Kanna

Bratati Maity

Deepika Gujjarlapudi

Sadhana Yelamanchili

