## Supplementary material for "Clinical outcomes in vaccinated individuals hospitalized with Delta variant of SARS-CoV-2": Figure S1

* Equal Contribution

### Corresponding authors

### Figure S1

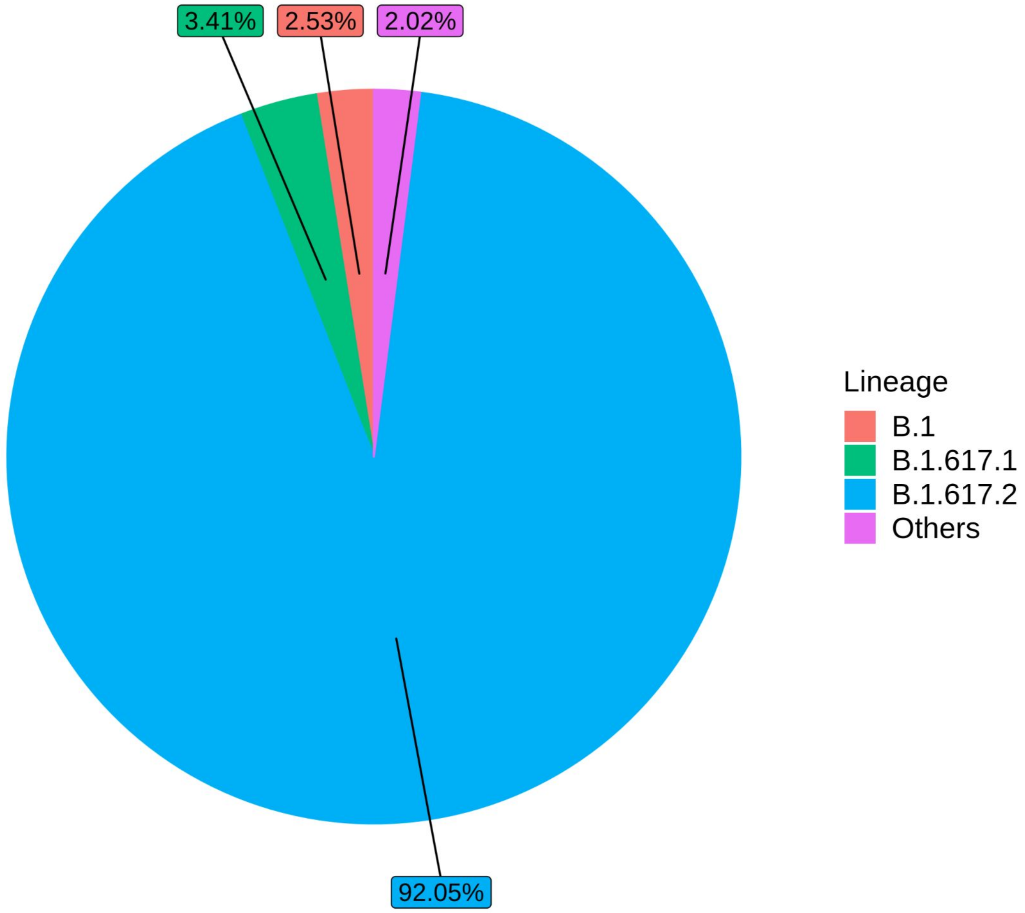


Lineages prevalent in community samples of Telangana, India, in the month of May 2021. Data source: GISAID. Only those genomes which were at least 95% covered are represented (n = 792).

### Figure S2

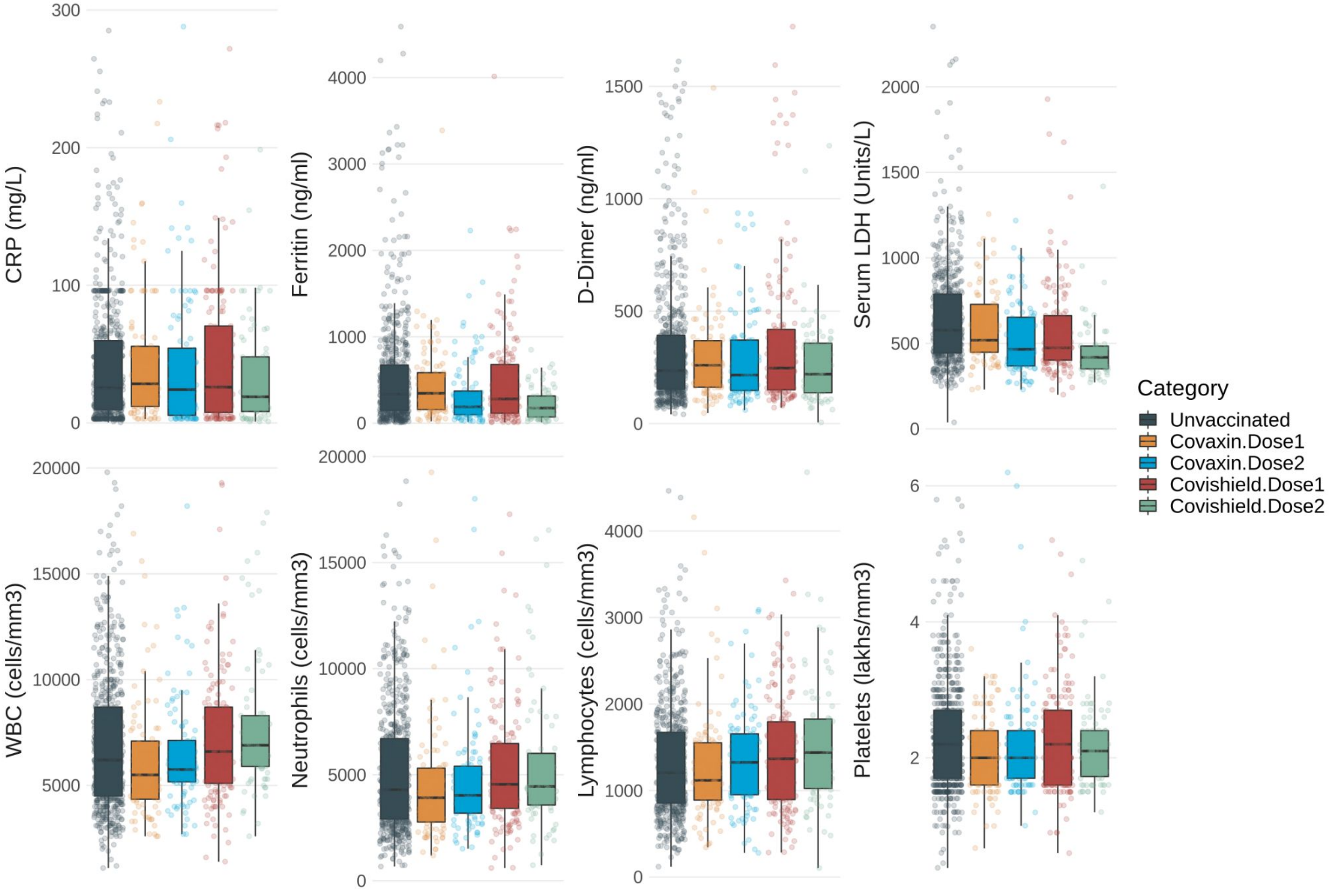


Inflammatory markers in vaccinated and unvaccinated individuals, grouped by vaccine type and number of doses.
